# Effect of a simple exercise programme on hospitalisation-associated disability in older patients: a randomised controlled trial

**DOI:** 10.1101/19008151

**Authors:** Javier Ortiz-Alonso, Natalia Bustamante-Ara, Pedro L. Valenzuela, María T. Vidán, Gabriel Rodríguez-Romo, Jennifer Mayordomo-Cava, Marianna Javier-González, Mercedes Hidalgo-Gamarra, Myriel Lopéz-Tatis, María Isabel Valades-Malagón, Alejandro Santos-Lozano, Alejandro Lucia, José Antonio Serra-Rexach

## Abstract

**Objective:** Hospitalisation-associated disability (HAD, defined as the loss of ability to perform one or more basic activities of daily living [ADL] independently at discharge) is a frequent condition among older patients. The present study aimed to assess whether a simple inpatient exercise programme decreases the incidence of HAD in acutely hospitalised very old patients.

**Design:** In this randomized controlled trial (Activity in GEriatric acute CARe, AGECAR) participants were assigned to a control or intervention (exercise) group, and were assessed at baseline, admission, discharge, and 3 months thereafter.

**Setting and participants:** 268 patients (mean age 88 years, range 75–102) admitted to an acute care for elders (ACE) unit of a Public Hospital were randomized to a control (n=125) or intervention (exercise) group (n=143).

**Methods:** Both groups received usual care, and patients in the intervention group also performed simple supervised exercises (walking and rising from a chair, for a total daily duration of ∼20 min). We measured incident HAD at discharge and after 3 months (primary outcome); and Short Physical Performance Battery (SPPB), ambulatory capacity, number of falls, re-hospitalisation and death during a 3-month follow-up (secondary outcomes).

**Results:** Median duration of hospitalisation was 7 days (interquartile range 4 days). Compared with admission, the intervention group had a lower risk of HAD at discharge (odds ratio [OR]: 0.32; 95% confidence interval [CI]: 0.11–0.92) and at 3-months follow-up (OR 0.24; 95% CI: 0.08–0.74) than controls during follow-up. No intervention effect was noted for the other secondary endpoints (all p>0.05), although a trend towards a lower mortality risk was observed in the intervention group (p=0.078).

**Conclusion and implications:** These findings demonstrate that a simple inpatient exercise programme significantly decreases the risk of HAD in acutely hospitalised, very old patients.

**Trial registration:** NCT0137489 (https://clinicaltrials.gov/ct2/show/NCT01374893).

**Brief summary:** A simple inpatient intervention consisting of walking and rising from a chair (∼20 minutes/day) considerably decreases the risk of hospitalisation-associated disability in acutely hospitalised older patients.

## INTRODUCTION

Hospitalisation-associated disability (HAD, *i*.*e*., loss of the ability to perform one or more basic activities of daily living [ADL] independently at discharge) occurs in more than one-third of hospitalised older adults.^1–4^ This condition is associated with long-term disability, institutionalisation and death,^5–7^ and therefore its prevention should be a priority.

Hospitalised older patients spend most of the time in bed even if they are able to walk independently,^8^ and the majority (73%) do not walk at all during hospitalisation.^9^ Several physical strategies can be applied to prevent the functional decline associated with periods of restricted mobility such as those imposed by hospitalisation, but physical exercise appears the most effective.^10^ Exercise interventions have proven feasible and safe in acutely hospitalised older medical patients, and are effective to improve their functional status at discharge as well as to reduce the length and cost of hospital stays.^11–13^ It has been recommended that hospitalised old people should perform multicomponent exercise programmes, that is, combining walking and resistance exercises.^11^ However, this type of intervention usually involves complex exercises and requires the purchase of weight-training equipment, which might hinder its routine implementation in some Acute Care of Elderly (ACE) units.

The aim of this study was to determine the effects of an inpatient exercise programme including simple physical exercises for patients admitted to an ACE unit on: incident HAD at discharge and at 3-months post-discharge (*primary endpoint*); and performance on the Short Physical Performance Battery (SPPB), independent-walking ability (functional ambulatory classification [FAC]), and number of falls, re-hospitalisation and mortality risk during the 3 months following discharge (*secondary endpoints*).

## METHODS

### Study design

This clinical trial (NCT01374893) complied with the recommendations of the Consolidated Standards of Reporting Trials (CONSORT) statement.^14^ Patients (>75 years) admitted to our ACE unit (*Hospital General Universitario Gregorio Marañón*, Madrid) from June 2012 to June 2014 were considered eligible to participate in the study. During these two years, the patients were eligible to be included in the control or intervention group in a time-dependent manner (i.e., using 4- or 8-week blocks), to avoid the co-presence of patients from both groups in the unit, such that the patients were blinded to actual group assignment. We then excluded those who were non-ambulatory or dependent in all basic ADL at baseline (i.e., 2 weeks before admission, assessed by retrospective interview), had unstable cardiovascular disease or any other major medical condition contraindicating exercise, terminal illness, severe dementia, an expected length of hospitalisation <3 days, were transferred from another hospital unit, or had a scheduled admission (which was usually associated with a length of hospitalisation <3 days). Only those hospitalised for 3 or more days and alive at discharge were included. The Institutional Review Board approved the protocol, and written informed consent was obtained from patients. When it was not possible to obtain the informed consent directly from a patient due to medical reasons (e.g., impaired cognitive function), proxy consent was obtained from their relatives. All procedures were performed in accordance with the ethical standards laid down in the 1964 Declaration of Helsinki and its later amendments. Both groups received standard care in our ACE unit.^15^

### Intervention

The intervention (Monday to Friday) consisted solely of rising from a seated to an upright position (using armrests/assistance if necessary) and supervised walking exercises (a representative example of the exercises performed is available as **Supplementary Material**). For the former, exercise loads increased individually and progressively from 1 to 3 sets of up to 10 repetitions, with a 2-minute rest between sets. When patients could complete the prescribed training session (e.g., 1 set of 10 repetitions) in two consecutive days, a new set was added, up to a maximum of 3 sets of 10 repetitions per session. Walking total time progressed from 3 to 10 minutes (with resting periods if needed, depending on the patient’s condition) along the corridor of the ward with/without assistance. Standing and walking exercises were separated by a rest period of up to 5 minutes. All the exercise sessions were supervised by a fitness specialist (NBA or GRR), and training loads were recorded in a notebook.

### Endpoints

The research staff included nurses, medical residents and the aforementioned fitness specialists who were in charge of supervising each session. Those involved in outcome assessment were not involved in supervising the intervention. However, assessors and care providers were not blinded to the assigned intervention. The ability to perform basic ADL, the SPPB,^16^ and the modified FAC^17^ were assessed in-hospital on admission and on discharge. Basic ADLs and FAC were also assessed retrospectively to analyse baseline status, as well as by telephone interview 3-months post-discharge. HAD was considered as a dichotomous variable attending to whether the patient had lost or not the ability to perform one or more ADL independently, and was assessed at discharge and 3 months later.^1^ The basic ADL included for HAD assessment were bathing, dressing, toileting, transferring, continence and eating.^18^ We also assessed the loss of independent ambulation at discharge and three months later. This was determined according to the modified FAC, which classifies patients in five different categories attending to their level of dependence during walking: a score of 0 was assigned if the patient could not walk, 1 if the patient required continuous manual contact to support the body, 2 for light or intermittent manual contact to assist balance, 3 for independent but supervised ambulation, and 4 for independent ambulation on level surfaces or stairs.^17^ Thus, a FAC score <4 was considered the threshold for independent ambulation.^17^ Mortality and number of falls during post-discharge follow-up were recorded by telephone interview.

Although the original protocol specified a change in SPPB score as the primary endpoint,^14^ an interim analysis after 4 months revealed that 94% (intervention) and 84% (controls) of patients had an SPPB score <6 on a 0–12 scale, masking a potential intervention effect. Accordingly, we considered that the intervention effects would not be noticeable in a test such as the SPPB given the very low physical performance capacity of the study participants, whereas ability to perform basic ADL might be more responsive to the intervention in this patient group. Thus, HAD was established as the primary endpoint.

### Statistical analysis

Prior (unpublished) data obtained in 604 patients showed that 33% of the patients admitted to our ACE unit improved in one or more ADL from admission to discharge. As we hoped to increase this rate by 15% with the intervention (up to 48%), we estimated a sample of 260 or more patients to detect a significant intervention effect on incident HAD (power=80%, α=0.05).

Data are presented as mean±standard deviation (SD). Normal distribution (Shapiro-Wilk test) and homoscedasticity (Levene’s test) of the data were checked before any statistical treatment. Groups were compared using Student’s independent or chi-square tests for continuous or dichotomous variables, respectively. Binary logistic regression was used to compare the risk of HAD, frailty (SPPB score <9), independent ambulation, re-hospitalisation and death between groups. Survival analysis (crude and adjusted Kaplan-Meier) was performed during the 3-month follow-up. Incidence rate ratio (IRR, calculated with negative binomial regression) was used to compare the number of falls between groups. We did not impute missing data, and thus only available data was used for analysis for each specific variable. All statistical analyses were conducted using a statistical software package (SPSS 23.0, IBM, NY) with α=0.05.

## RESULTS

A flow diagram of study participants is shown in **figure 1**. From a total of 1110 patients admitted to our ACE unit during the study period, 281 met all inclusion criteria and volunteered to participate. Of these, 131 and 150 were assigned to the control and the intervention group, respectively. Finally, 268 patients (n=143 and 125 for intervention and control, respectively) completed the study and could be included in the analysis. No between-group differences were found at the start of the study for most sociodemographic/clinical variables, but those in the intervention arm were overall more dependent in ADL function, both at baseline (i.e., 2 weeks before admission, assessed by retrospective interview) and admission (**Table 1**). The median length of hospitalisation was 7 days (interquartile range [IQR] 4), with no between-group differences (p=0.246). Median duration of the exercise intervention was 3 days (IQR 2), with an average total exercise time per day of ∼20 min (the median duration of the walking part was 5 min [IQR 4, range 0 to 10] and patients performed a mean of 9 [SD 6, range 0 to 30] sit to stands). No adverse effects or falls were recorded during the intervention. Five (3.5%) patients did not exercise because they had a rapid, severe functional decline shortly after admission.

**Table 1.** Baseline, admission and main inhospital characteristics by group

| Variable | Control<br>(N=125) | Exercise<br>(N=143) | p-<br>value |
| --- | --- | --- | --- |
| Age (mean ± SD), years | 88 ± 5 | 88 ± 5 | 0.88 |
| Women | 54% | 60% | 0.28 |
| Body mass index (mean ± SD), kg/m<sup>2</sup> | 26.0 ± 6.4 | 26.1 ± 9.3 | 0.95 |
| Living at home | 93% | 91% | 0.57 |
| Charlson comorbidity index (mean ± SD) | 6.8 ± 1.9 | 6.7 ± 1.7 | 0.88 |
| **Geriatric syndromes** |  |  |  |
| Dementia | 12% | 27% | **0.003** |
| Depression | 18% | 32% | **0.010** |
| Falls | 16% | 36% | **<0.001** |
| Chronic pain | 30% | 35% | 0.43 |
| Malnutrition | 14% | 22% | 0.085 |
| Urinary incontinence | 39% | 49% | 0.11 |
| Frailty phenotype <sup>a</sup> | 65% | 74% | 0.097 |
| Incident delirium | 18% | 28% | 0.81 |
| **Polypharmacy ($\geq 7$)** | 55% | 58% | 0.64 |
| **Main admission diagnosis** |  |  | 0.39 |
| Respiratory | 26% | 34% |  |
| Circulatory | 30% | 26% |  |
| Renal/urologic | 15% | 8% |  |
| Central nervous system | 7% | 13% |  |
| Digestive | 7% | 8% |  |
| **Independence in basic ADLs at baseline<sup>b</sup>** |  |  |  |
| Full function ($>4$ ADLs) | 62% | 50% | **0.036** |
| Moderate impairment (3–4 ADLs) | 22% | 24% | 0.58 |
| Severe impairment ($<3$ ADLs) | 16% | 26% | **0.049** |
| **Independent ambulation at baseline (FAC $<4$)** | 76% | 68% | 0.14 |
| **Independence in basic ADLs at admission<sup>b</sup>** |  |  |  |
| Full function ($>4$ ADLs) | 30% | 18% | **0.019** |
| Moderate impairment (3–4 ADLs) | 25% | 21% | 0.46 |
| Severe impairment ($<3$ ADLs) | 45% | 61% | **0.009** |
| **Independent ambulation (FAC $<4$) at admission** | 40% | 31% | 0.13 |
| **SPPB (mean ±SD), score at admission** | 3.8 ± 2.9 | 3.2 ± 2.5 | 0.078 |
| **Length of hospitalization (median [IQR]), days** | 7 (5) | 6 (4) | 0.25 |

Abbreviations: ADLs, activities of daily living; FAC, functional ambulation category; IQR, interquartile range; SPPB, short physical performance battery.
<sup>a</sup> Frailty was defined as having $\geq 3$ of 5 Fried's criteria.<sup>26</sup>
<sup>b</sup> The 6 basic ADLs assessed were bathing, dressing, transferring, toileting, continence and feeding. Significant differences between groups ( $p < 0.05$ ) are in bold.

**Figure 1.**
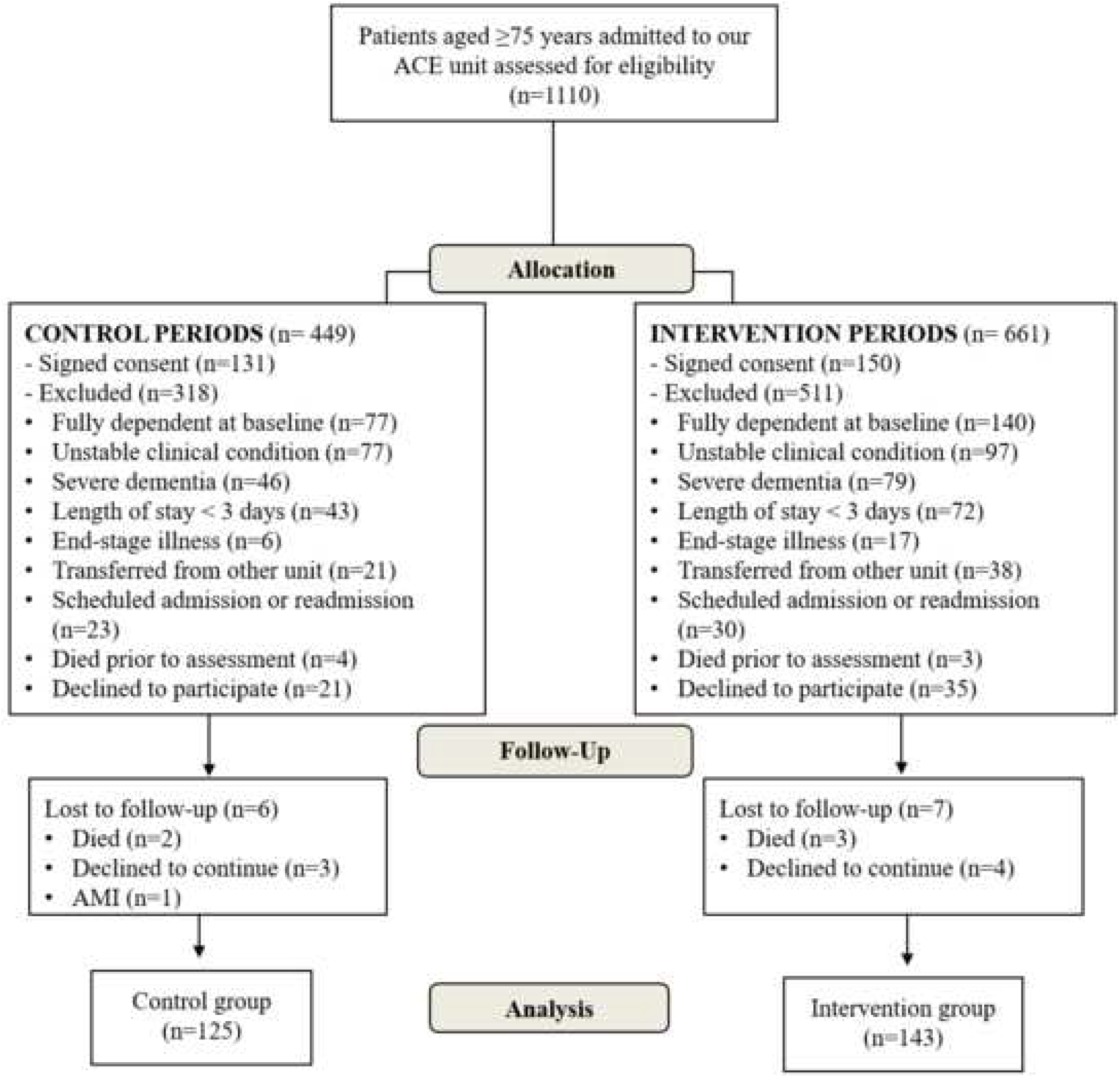
Flowchart of study participants. Abbreviations: AMI, acute myocardial infarction.

Compared with admission, patients in the exercise group had a significantly lower risk of HAD than their controls both at discharge and at 3-months post-discharge, even when adjusting for clinical characteristics and functional performance at admission (**Table 2**), with the proportion of patients diagnosed with HAD significantly lower in the former at both time points (p=0.012 and 0.003 for discharge and 3-month post-discharge, respectively). Compared with baseline, however, the proportion of patients with HAD did not differ between groups at discharge (56% and 58% for the intervention and control group, respectively; p=0.685) or 3 months later (53% and 62%, respectively; p=0.188). Also, no intervention effect was noted at discharge for the SPPB (3.6±2.5 and 4.2±3.1 for the intervention and control group, respectively; p=0.108) and FAC score (2.8±1.2 and 2.9±1.3, respectively; p=0.404), or for risk of re-hospitalisation or number of falls during the 3-month follow-up (0.44±1.47 [median: 0, range: 0, 10] and 0.70±1.70 [median: 0, range: 0, 10]) (**Table 2**). A non-significant trend towards a lower mortality risk was observed in the intervention group, with crude and adjusted Kaplan-Meier analysis confirming this trend (p=0.084 and 0.085, respectively) (**figure 2**).

**Table 2.**
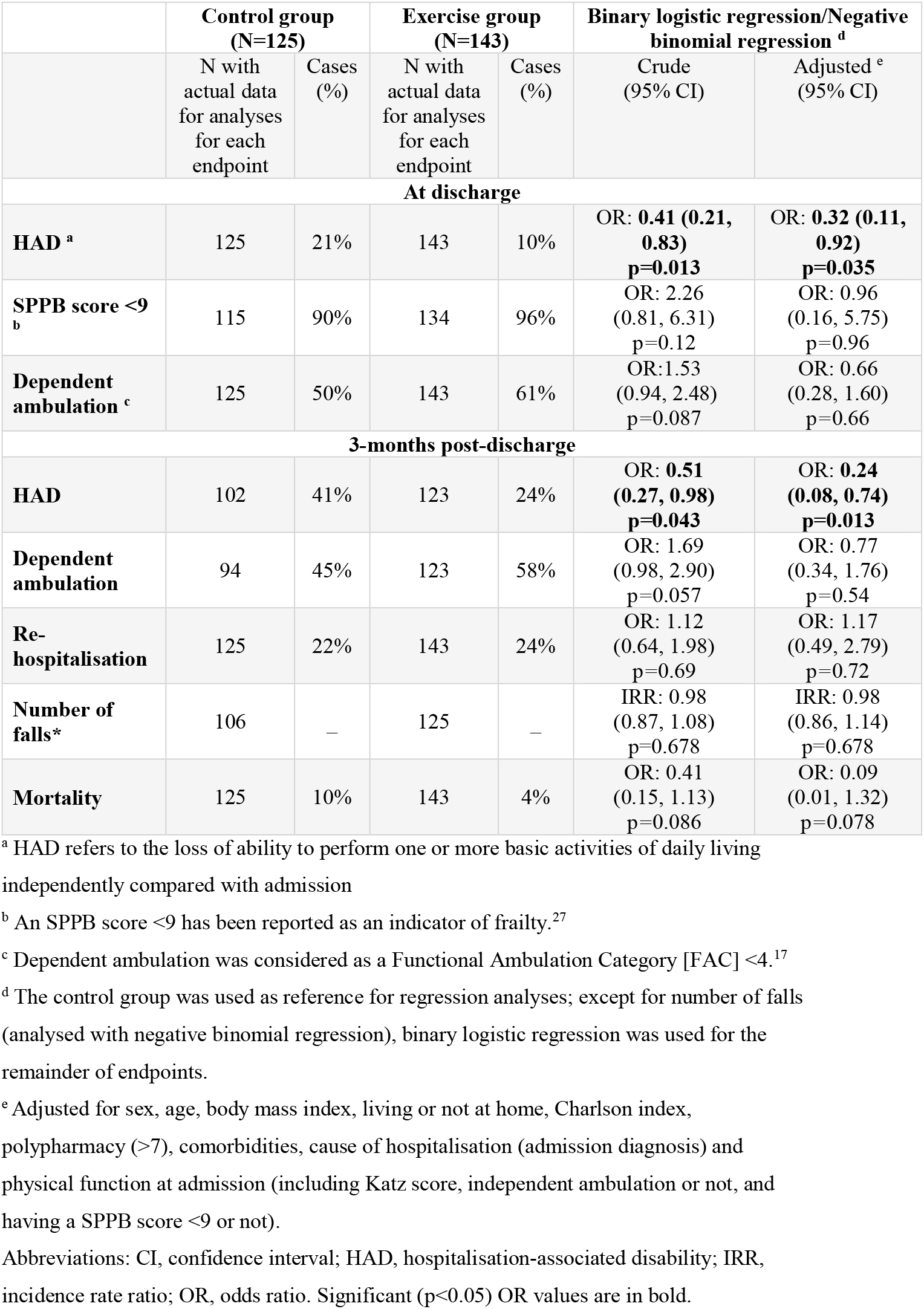
Effects of hospitalization on study endpoints

**Figure 2.**
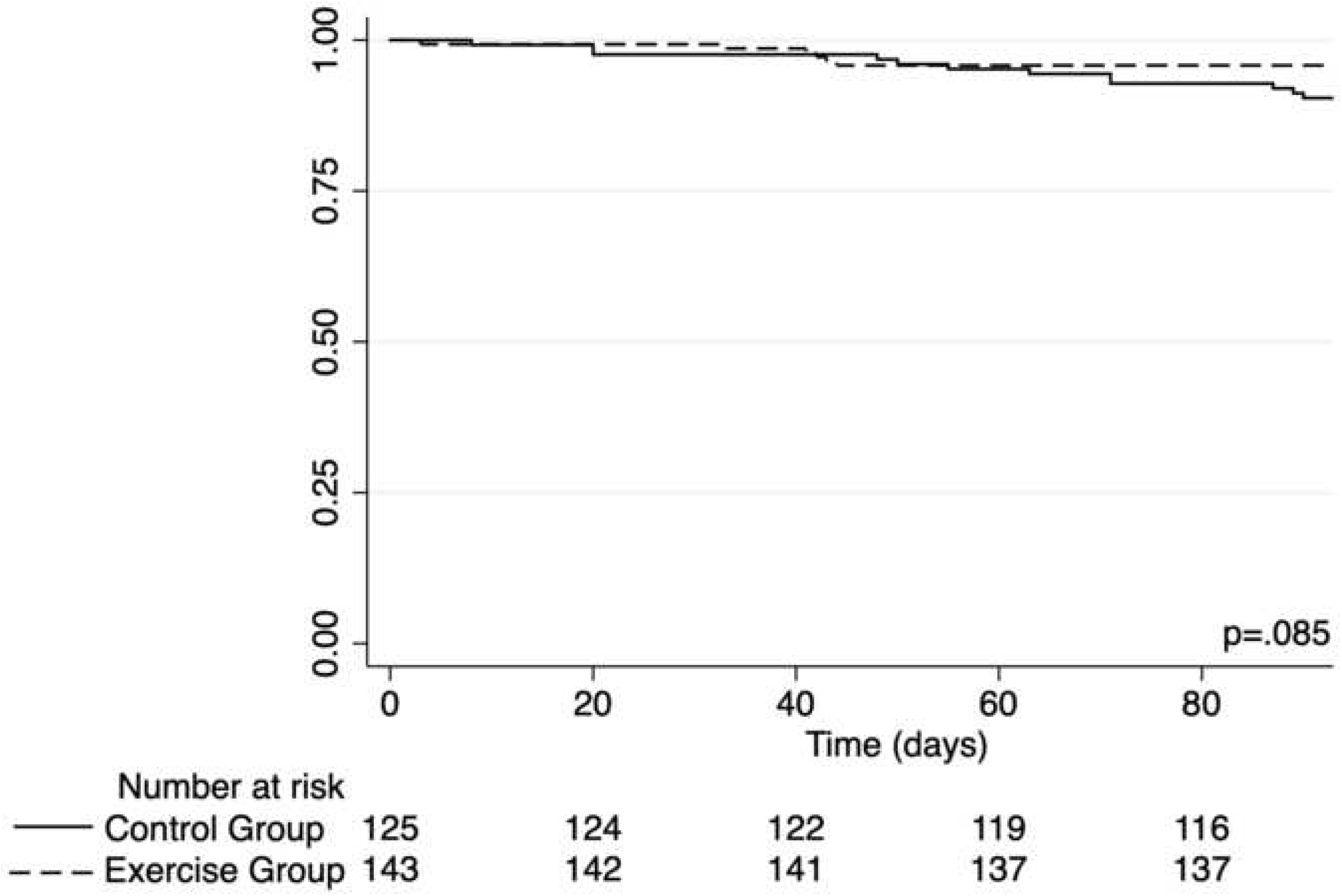
Kaplan-Meier survival analysis.

## DISCUSSION

HAD is a frequent condition among hospitalised older adults that has major negative consequences for patients and caregivers.^1^ Our finding that performing simple inpatient exercises (solely walking and rising from a chair) considerably decreases (by ∼70%) the risk of HAD in acutely hospitalised patients of advanced age (mean of 88 years), the majority also being frail, is thus important. Accordingly, our results support the routine implementation of these exercises during the acute hospitalisation of older patients.

Results of different systematic reviews indicate that inpatient exercise programmes are safe and overall effective in hospitalised older patients.^11–13^ However, not all types of exercise interventions are easily feasible in this population owing to a frequently limited mobility capacity. Walking during hospital stay has proven to maintain pre-hospitalisation mobility, but conflicting results have been reported regarding its benefits on ADL function.^19,20^ By contrast, the combination of walking with lower limb strengthening exercises can provide greater benefits on functional ability.^21,22^ Indeed, walking combined with stretching/strengthening exercises (*e*.*g*., leg lifts/swings, toe/heel raises) is associated with a better ADL function at 1-month post-discharge.^21^ A recent study reported that a combination of walking, balance, and resistance exercises during a median of 5 days provided significant benefits in the functional ability of older patients during acute hospitalisation.^22^ But, the aforementioned intervention was more complex than the present protocol and required specific weight-training equipment,^22^ which might hinder routine implementation in some ACE units.

Although sedentary behaviours (such as those commonly observed in hospitalised patients ^8^) are associated with an increased incidence of frailty in older adults, the inclusion of a low amount of physical activity moderates (or even completely offsets if performed for >27 minutes) this relationship,^23^ which reinforces the need of increasing physical activity levels regardless of the specific exercise performed. In further support of the effectiveness of simple exercises against sedentary behaviours and the consequent functional decline, Harvey et al. recently reported that encouraging frequent but small bouts of exercise (i.e., rising from a chair) might suffice to reduce functional decline in frail sheltered housing residents.^24^ It is reasonable to hypothesise that, as in healthy older adults, a dose-response relationship likely exists between training intensity and exercise benefits.^25^ However, our results suggest that walking for up to 10 minutes and standing up from a chair (gradually increasing exercise loads by increasing the total number of repetitions [until a maximum of 30/day] and providing external help during the movement if needed) might induce clinically meaningful benefits in the case of frail older patients such as those studied here. That said, this type of intervention requires close supervision and thus an additional time involvement of the hospital staff with respect to their daily duties or reliance on external staff, as described here (i.e., fitness specialists).

Several potential limitations should be considered. Groups initially differed in functional and mental status as well as in falls history, which might have potentially biased the outcomes. In this respect, however, the benefits of the intervention on the risk of incident HAD remained significant even after adjusting for clinical and functional variables at admission. The losses during the follow-up (up to ∼20% of participants for some endpoints such as FAC) should also be borne in mind as a limitation. Also, our study is a single-centre study performed in a very old and frail population, which could limit the generalisability of our findings to younger or more functional patients. The small proportion (∼25%) of the total number of hospitalised patients in our ACE unit that participated in our study possibly (partly due to the strict inclusion criteria) lends further support to the idea that the present results might not be necessarily applicable to acutely hospitalised old patients in general. On the other hand, although this intervention did not require investment in weight-training equipment, the additional time commitment needed from the hospital staff to supervise the exercise sessions (or alternatively the need to rely on external staff such as fitness specialists, as we did here) are issues that should be addressed in future research, for instance in studies aiming to assess the cost effectiveness of the present type of intervention. It must also be noted that no intervention benefits were observed when HAD was analysed with respect to baseline (i.e., 2 weeks before admission). Finally, the greater functional independence of the control group at baseline could have potentially influenced the results, as the patients of this group might have tended to more rapidly recover their “normal” physical status during hospitalisation even in the absence of an exercise intervention.

## CONCLUSIONS AND IMPLICATIONS

A simple inpatient exercise programme solely consisting of walking and rising from a chair (median duration of the intervention only 3 days, ∼20 minutes/day) decreases the incidence of HAD in acutely hospitalised, very old patients. Further research is needed to analyse the cost-effectiveness of this type of intervention (in terms of staff resources) and its generalisability. However, given the clinical relevance of HAD, our results support the routine implementation of these exercises during the acute hospitalisation of older patients.

## Data Availability

Data will be made available upon request to the corresponding author.

## Acknowledgements

We sincerely thank the geriatrics ward team for their valuable help and contribution to our results, with a special mention to the following professionals: Alison Martínez, Jaqueline Ferrero, Mar Alonso, Laura Arribas, Laura Osuna, Reyes Vaquerizo, Cristina Cebollero, Yolanda Navarro, Soledad Rubio, Ana Ruiz, Saray Martínez, Raquel Moreno, María Angeles Flores, Lidia del Campo, Azucena Ríos, Antonio Piñeiro, Manuela Gámez, Piedad Trujillo, Mayte Mesa, Raquel Gutiérrez, Ana M^a^ García, M^a^ Jesús Ruiz, Susana Palacios, Marta Orea, Ana Belén Huguet, Laura Jaro, Susana Robledo, José Lázaro.

